# Application of multilevel linear spline models for analysis of growth trajectories in a cohort with repeat antenatal and postnatal measures of growth: a prospective cohort study

**DOI:** 10.1101/2022.05.03.22274407

**Authors:** Linda M. O’Keeffe, Cara A. Yelverton, Helena C. Bartels, Kate N. O’Neill, Ciara McDonnell, Fionnuala M. McAuliffe

## Abstract

**Introduction:** Antenatal and postnatal growth are important indicators of fetal and child health and development. Studies frequently have repeat antenatal and postnatal measures of growth available and require approaches that can maximise the use of these measures to examine growth trajectories. We demonstrate the use of multilevel linear spline modelling to model growth trajectories with repeated antenatal and postnatal measures of growth from 20 weeks gestation to five years in a cohort at high risk of macrosomia.

**Methods:** Prospective follow-up data from 720-759 mother-child pairs from the ROLO study (initially a randomized controlled trial of a low glycemic index diet in pregnancy to prevent recurrence of macrosomia [birthweight > 4K]) were analysed. Fetal measurements were obtained from ultrasound scans performed on mothers at 20-and 34-weeks gestation, including abdominal circumference (AC) and head circumference (HC). An estimated fetal weight was obtained at 20-and 34-weeks gestation, calculated using the Hadlock 4-parameter formula. At delivery, AC, HC, weight and length were recorded. Follow-up anthropometry assessments (AC, HC, weight and length/height) were also obtained in childhood at six months, two years and five years. Linear spline multilevel models were used to examine trajectories of AC, HC and weight from 20 weeks gestation to five years and length/height from birth to five years.

**Results:** 754, 756 and 759 participants were included in analyses of AC, HC and weight respectively, while 720 participants were included in analysis of length/height. Over 50% of women had 3^rd^ level education and over 90% were of White ethnicity. Women were a mean (SD) age of 32 (4.2) at recruitment. Following exploration of a series of different models for each growth measure, the best fitting model for AC, HC and weight included a model with knots at each measurement occasion giving rise to five linear spline periods from: 20 weeks to 34 weeks gestation, 34 weeks gestation to birth, birth to six months, six months to two years and two years to five years. The best fitting models for length/height included a model with three linear spline periods from birth to six months, six months to two years and two years to five years. Comparison of observed and predicted values for each model demonstrated good model fit. For all growth measures, fetal growth rates were generally fastest in pregnancy or immediately postpartum (for length/height), with rates of growth slowing after birth and becoming slower still as infancy and childhood progressed. We found little difference in growth trajectories between the intervention and control group. There was some evidence of slightly lower HC, weight and length among females compared with males at birth which appeared to widen by age five years due to slower postnatal growth rates among females.

**Conclusion:** We demonstrate the application of multilevel linear spline models for examining growth trajectories when both antenatal and postnatal measures of growth are available. The approach may be useful for cohort studies or randomised controlled trials with repeat prospective assessments of growth spanning pregnancy and childhood.

## Introduction

Antenatal and childhood growth are important indicators of fetal and child health and development and are associated with health in adult life (1, 2). Consequently, modelling of growth trajectories, identifying causes and predictors of different growth trajectories and relating growth trajectories in the early life course to later life health is important for informing a life course approach to disease prevention (3-5).

A key aspect of understanding growth patterns, their causes, predictors and outcomes includes appropriate modelling of longitudinal growth data (3). Since repeated measures of growth within individuals are not independent of each other and the scale and variance of growth measures often changes over time, traditional approaches to analysis of growth data, such as Z-score based methods analysed using multiple regression, do not take account of the clustering of repeated measures within individuals (3). Moreover, the true shape of growth trajectories cannot be modelled using such approaches. While appropriate methods for the study of longitudinal growth data have been applied to antenatal and childhood growth measures in many cohort studies, most studies to date have examined antenatal growth (6, 7) or postnatal growth as separate processes/trajectories (8-14). Appropriate modelling of growth data as a continuum from antenatal to postnatal life is important to accurately characterise the shape of growth from early gestation into childhood to better understand it’s aetiology. In addition, it also allows such trajectories to be examined as outcomes for pre-conception or early pregnancy exposures or to be examined themselves as exposures for later health outcomes (3).

Using data from the prospective follow-up of a randomised controlled trial of a low glycaemic index diet in pregnancy (ROLO study), we demonstrate the application of linear spline multilevel models for modelling antenatal and postnatal growth trajectories using four measures of anthropometry (abdominal circumference [AC], head circumference [HC], weight and length/height) from 20 weeks’ gestation to age five years.

## Methods

### Study population

The ROLO study is a randomised control trial of a low glycaemic index diet in pregnancy that recruited 800 secundigravid women who had previously given birth to a baby weighing over 4kg between 2007-2011 at the National Maternity Hospital, Dublin, Ireland (15). Women were recruited at first antenatal consultation. Women with any underlying medical disorders, including a previous history of gestational diabetes, those on any drugs, those unable to give full informed consent, aged less than 18 years, of gestation greater than 18 weeks, and having multiple pregnancies were excluded. Women were randomised to either the intervention group which received dietary advice on a low glycaemic diet, or the control group who received routine antenatal care.

### Measurement of anthropometry

#### Antenatal measures

Fetal measurements were obtained from ultrasound scans performed on mothers at medians of 20 + 6 (Interquartile Range [IQR]: 20 + 1 to 21 + 5) and 34 + 1 (IQR: 33 + 5 to 34 + 5) weeks’ gestation, including AC and HC. An estimated fetal weight (EFW) at 20- and 34-weeks’ gestation was calculated using the Hadlock 4-parameter formula. Ultrasound measurements were taken by two ultrasonographers using a Voluson 730 Expert (GE Medical Systems, Germany) using standard procedures.

#### Postnatal measures

At delivery, infants’ AC, HC, weight and length were recorded. Follow-up anthropometry assessments were also obtained in childhood at six months, two years and five years (15-17). All measurements were obtained and calculated by a trained member of the research team. At six months, two years and five years, weight (kg) of the child was measured using a calibrated stand on digital weighing scale (SECA 813) to the nearest 0.1 kg by a trained research team member. Children were measured in light clothing without shoes. Standing height was measured, without shoes, with head aligned in the Frankfort plain, using a free-standing stadiometer (SECA 217) and measurements recorded to the nearest 0.1cm. The child’s head and abdominal circumferences were measured using a SECA ergonomic circumference measuring tape, to the nearest 0.1cm. All measurements were recorded three times and the average calculated to improve reliability.

### Statistical analysis

We used multilevel models to examine trajectories of change in AC, HC, weight and length/height from 20 weeks gestation to age five years (18, 19). Multilevel models estimate mean trajectories of the outcome while accounting for the non-independence (i.e. clustering) of repeated measurements within individuals, change in scale and variance of measures over time and differences in the number and timing of measurements between individuals (using all available data from all eligible participants under a Missing at Random [MAR] assumption) (3, 20).

Change in all four growth measures was estimated here using linear spline multilevel models (two levels: measurement occasion and individual) (3). Linear splines allow knot points to be fit at different ages to derive periods in which change is approximately linear. The optimal linear spline model for each growth measure was selected by examining observed data for each growth measure and comparing model fit statistics of different models including models that assumed linear change over time to models with knot points at different ages (strategies for selection of knot points are described elsewhere in detail (3)). Model fit statistics examined included Akaike’s Information Criterion and observed and predicted values of each growth measure across the age range of the model. Following exploration of a series of models, the best fitting model for AC, HC and weight included a model with knots at each measurement occasion giving rise to five linear spline periods from 20 weeks’ to 34 weeks’ gestation, 34 weeks’ gestation to birth, birth to six months, six months to two years and two years to five years while the best fitting models for length/height included a model with three linear spline periods from birth to six months, six months to two years and two years to five years.

All outcomes were normally distributed at each measurement occasion. Except for length/height which did not include antenatal measures, trajectories were centred on the first available measure (20 weeks gestation) for AC, HC and weight. Length/height trajectories were centred at birth. For all models we placed no restrictions on the variance-covariance matrices of level two (individual level) random effects. Given the substantial change in scale and variance of growth from antenatal to postnatal life, we also aimed to allow occasion level measurement error to vary with age (level one random effects for the slope). Therefore, all models included a level one random effect for the slope while the HC model also included a level one random effect for the intercept. The final models for growth trajectories from 20 weeks gestation took the following form: AC_ij_ / HC_ij_ /weight_ij_= β_0_ + u_0j_ + (β_1_+ u_1j_)s_ij1_ + (β_2_+ u_2j_)s_ij2_ + (β_3_ + u_3j_)s_ij3_ + (β_4_ + u_4j_)s_ij4_ + (β_5_ + u_5j_)s_ij5_ + e_ij_ where for person j at measurement occasion i; β_0_ represents the fixed effect coefficient for the average intercept, β_1_ to β_5_ represent fixed effect coefficients for the average linear slopes of each linear spline, u_0j_ to u_5j_ indicate person-specific random effects for the intercept and slopes respectively, and e_ij_ represents the occasion-specific residuals or measurement error which were allowed to vary with age. The final model for length took a similar form but with only three linear spline periods due to the absence of measures prior to birth. Code for the application of these models using the “runmlwin” command from MlWin (21) within Stata 16 (22) is included in Supplementary Material.

## Results

754, 756 and 759 offspring were included in analyses of AC, HC, and weight respectively while 720 offspring were included in analyses of length/height. Table 1 includes the number of measures of each growth measure at each measurement occasion with number of measures available broadly similar across growth measures; for example, weight measures available on each occasion included 655 measures at 20 weeks gestation, 730 at 34 weeks gestation, 756 at birth, 280 at six months, 339 at two years and 387 at five years.

**Table 1.** N repeated measures included in analyses for each growth measure.

|  | **20 weeks** | **34 weeks** | **Birth** | **6 months** | **2 years** | **5 y ears** |
| --- | --- | --- | --- | --- | --- | --- |
| **Abdominal circumference** | 656 | 732 | 265 | 280 | 336 | 385 |
| **Head circumference** | 656 | 700 | 634 | 280 | 333 | 386 |
| **Weight** | 655 | 730 | 756 | 280 | 339 | 387 |
| **Length/height** |  |  | 634 | 280 | 339 | 386 |

Of participants included in analyses (Table 2), over 50% had completed third level education and a majority (>90%) were of White ethnicity. Among mothers of male babies, mean age (standard deviation (SD)) at delivery was approximately 32.3 (4.2) years, mean (SD) BMI at delivery was 27.1 (5.2) kg/m^2^, mean (SD) birthweight at delivery was 4.1 (0.5) kg and median (interquartile range (IQR)) gestational age was 40.4 (39.6, 41.1) weeks. Mothers of male babies had relatively low levels of deprivation as indicated by the mean (SD) Pobal HP index of 5.3 (10.8). Characteristics were broadly similar for mothers of female babies though mothers of female babies had somewhat higher levels of third level education (∼60%). Model fit as judged by differences between observed growth measures and those predicted by the models for AC, HC, weight and length are shown in Tables 3-6. Overall, our models have good model fit as all reference ranges for the difference between observed and predicted are less than the SD of the observed or less than 10% of the observed value which can be used as a rule of thumb for the assessment of model fit.

**Table 2.**
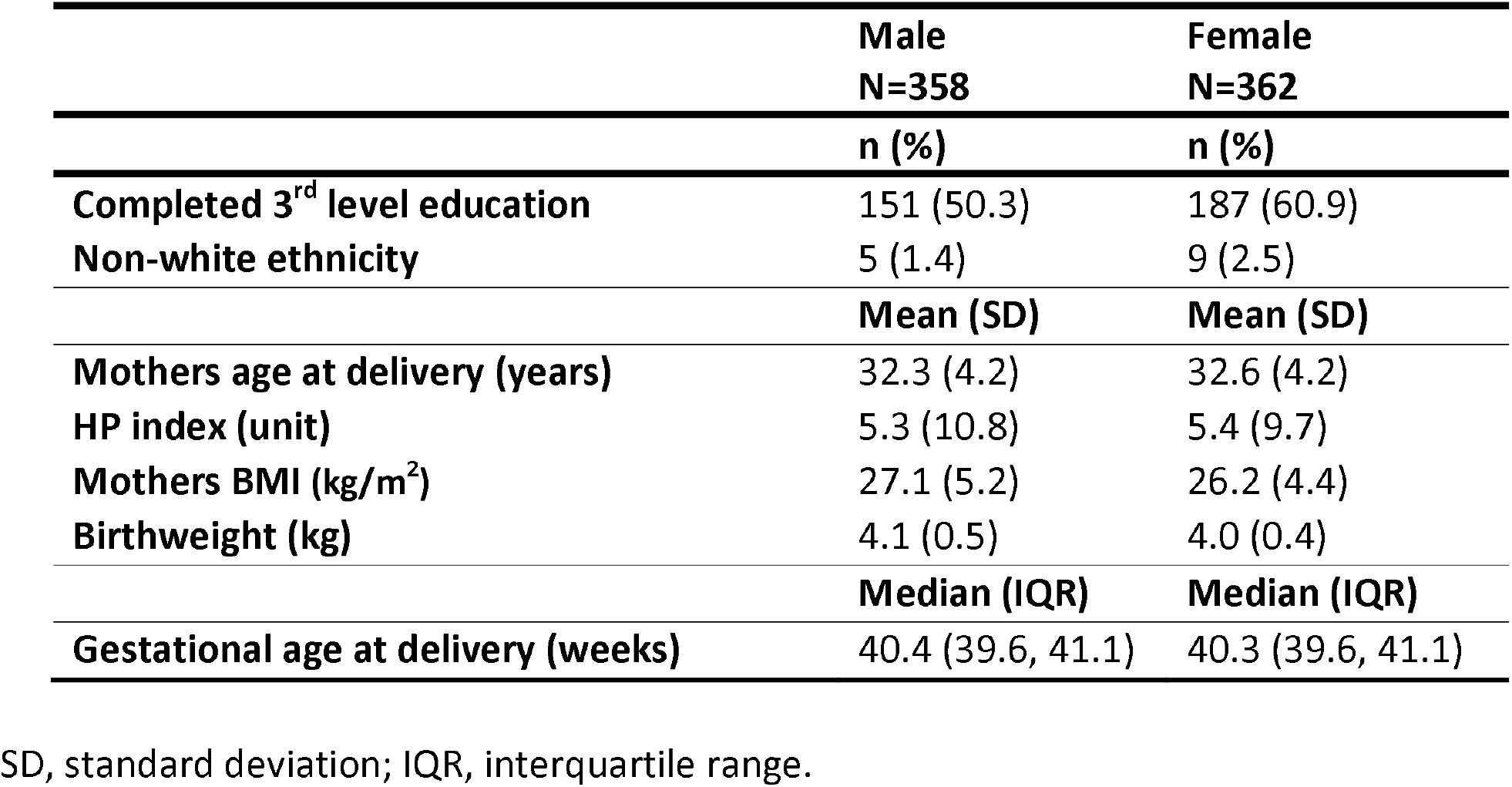
Characteristics of ROLO participants included in the analysis of length/height, by sex.

**Table 3.** Model details for abdominal circumference.

|  | **Total**<br>**number of**<br>**observations** | **Mean**<br>**observed**<br>**(SD) in cm** | **Mean**<br>**predicted**<br>**(SD) in cm** | **Mean**<br>**difference**<br>**(observed –**<br>**predicted) in cm** | **95% level of**<br>**agreement**<br>**between observed**<br>**and predicted in**<br>**cm** |
| --- | --- | --- | --- | --- | --- |
| **20 wks to 34 wks** | 531 | 22.48 (6.92) | 22.59 (6.86) | -0.08 | -1.13 to 0.97 |
| **34 wks to birth** | 517 | 32.22 (2.08) | 32.35 (1.19) | 0.08 | -2.01 to 2.17 |
| **Birth to 6 months** | 315 | 38.21 (5.93) | 36.01 (4.51) | -0.05 | -3.64 to 3.53 |
| **6 months to 2 years** | 272 | 47.91 (5.37) | 47.84 (3.96) | 0.09 | -4.25 to 4.42 |
| **2 years to 5 years** | 681 | 50.25 (8.98) | 54.03 (2.72) | -0.06 | -6.11 to 6.00 |

SD, standard deviation; wks, weeks.

**Table 4.** Model details for head circumference.

|  | **Total**<br>**number of**<br>**observations** | **Mean**<br>**observed**<br>**(SD) in cm** | **Mean**<br>**predicted**<br>**(SD) in cm** | **Mean**<br>**difference**<br>**(observed –**<br>**predicted) in cm** | **95% level of**<br>**agreement**<br>**between observed**<br>**and predicted in**<br>**cm** |
| --- | --- | --- | --- | --- | --- |
| **20 wks to 34 wks** | 292 | 22.90 (4.87) | 22.86 (4.83) | 0.09 | -0.55 to 0.73 |
| **34 wks to birth** | 680 | 32.54 (1.67) | 32.63 (1.53) | -0.01 | -0.61 to 0.59 |
| **Birth to 6 months** | 642 | 37.57 (3.70) | 37.39 (3.46) | 0.00 | -0.48 to 0.47 |
| **6 months to 2 years** | 274 | 47.29 (2.98) | 47.28 (2.87) | 0.01 | -0.47 to 0.49 |
| **2 years to 5 years** | 661 | 48.89 (6.16) | 51.18 (1.64) | -0.01 | -0.75 to 0.73 |

SD, standard deviation; wks, weeks.

**Table 5.**
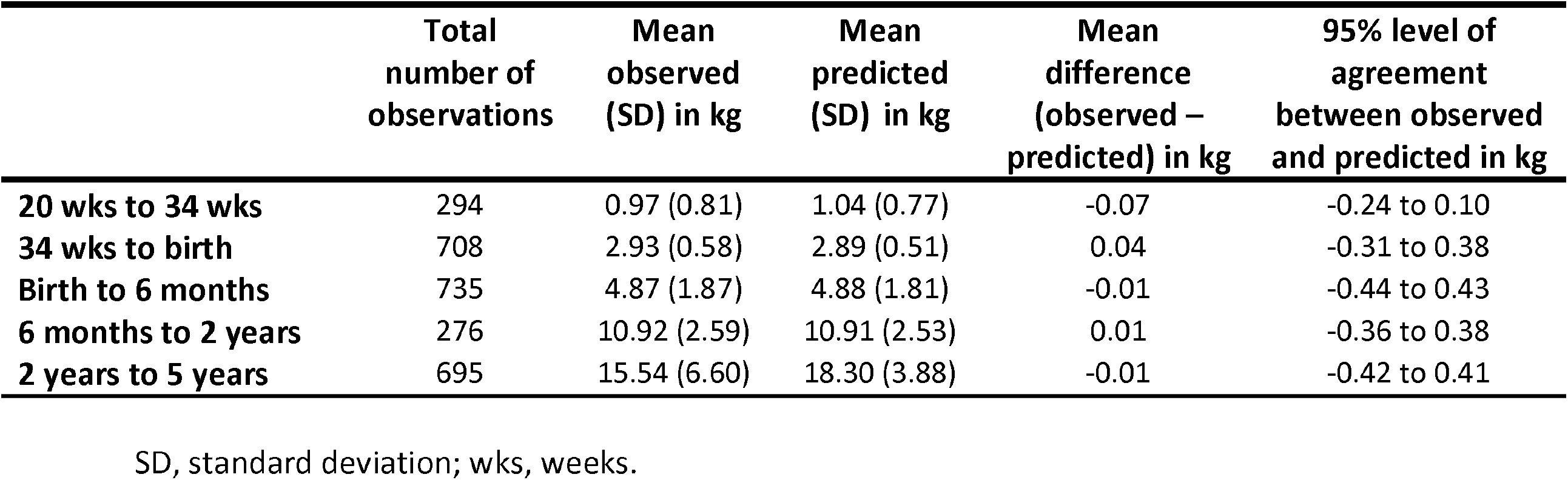
Model details for weight.

**Table 6.** Model details for length.

|  | **Total number**<br>**of**<br>**observations** | **Mean**<br>**observed**<br>**(SD) in cm** | **Mean**<br>**predicted**<br>**(SD) in cm** | **Mean difference**<br>**(observed –**<br>**predicted) in cm** | **95% level of**<br>**agreement**<br>**between**<br>**observed**<br>**and**<br>**predicted in**<br>**cm** |
| --- | --- | --- | --- | --- | --- |
| **Birth to 6 months** | 475 | 57.55 (7.51) | 57.14 (7.25) | 0.0001 | -0.03 to 0.03 |
| **6 months to 2 years** | 304 | 81.03 (10.12) | 81.03 (10.08) | -0.002 | -0.57 to 0.56 |
| **2 years to 5 years** | 574 | 104.07 (12.24) | 106.42 (9.79) | 0.0004 | -2.92 to 2.92 |

SD, standard deviation; wks, weeks.

Trajectories of AC, HC and weight from 20 weeks’ gestation to five years and trajectories of length/height from birth to five years by intervention status and sex are shown in Table 7 and Figures 1-4. AC and HC had the fastest rates of growth from 20 to 34 weeks’ gestation with growth rates continuing to slow thereafter up to age five years. Weight had the fastest growth rate from 34 weeks’ gestation to birth with growth rates slowing somewhat from birth to six months and continuing to slow thereafter until five years. Length/height had the fastest growth rates from birth to two years, with the growth rate decreasing thereafter and slowing further from two years to five years.

**Table 7.** Mean trajectories of anthropometry and mean difference in trajectories by intervention status and sex in the ROLO cohort.

|  | Mean trajectory (95% CI) in intervention | Mean difference in trajectory (95% CI) in controls | Mean trajectory (95% CI) in males | Mean difference in trajectory (95% CI) in females |
| --- | --- | --- | --- | --- |
| **Abdominal circumference** |  |  |  |  |
| 20 weeks (cm) | 15.96 (15.85,16.07) | -0.02 (-0.17,0.14) | 16.05 (15.94,16.16) | -0.20 (-0.35,-0.04) |
| 20 wks to 34 wks (cm/week)* | 1.20 (1.18,1.22) | 0.01 (-0.02,0.03) | 1.20 (1.19,1.22) | 0.002 (-0.02,0.03) |
| 34 wks to birth (cm/week)* | 0.26 (0.19,0.33) | 0.03 (-0.07,0.13) | 0.28 (0.21,0.35) | -0.01 (-0.11,0.09) |
| Birth (cm) | 34.31 (33.94,34.68) | 0.26 (-0.26,0.77) | 34.55 (34.18,34.93) | -0.22 (-0.74,0.29) |
| Birth to 6 months (cm/week)* | 0.40 (0.37,0.43) | -0.01 (-0.05,0.03) | 0.41 (0.38,0.44) | -0.03 (-0.06,0.01) |
| 6 months to 2 years (cm/week)* | 0.08 (0.07,0.09) | -0.001 (-0.01,0.01) | 0.07 (0.06,0.08) | 0.02 (0.004,0.03) |
| 2 years to 5 years (cm/week)* | 0.03 (0.02,0.03) | -0.0002 (-0.01,0.01) | 0.03 (0.02,0.03) | -0.002 (-0.01,0.01) |
| 5 years (cm) | 55.46 (54.91,56.02) | -0.03 (-0.82,0.76) | 55.33 (54.76,55.90) | 0.23 (-0.57,1.02) |
| **Head circumference** |  |  |  |  |
| 20 weeks (cm) | 18.60 (18.52,18.68) | -0.11 (-0.22,0.01) | 18.68 (18.60,18.76) | -0.27 (-0.38,-0.16) |
| 20 wks to 34 wks (cm/week)* | 1.01 (1.00,1.02) | -0.002 (-0.02,0.01) | 1.02 (1.01,1.03) | -0.01 (-0.02,0.004) |
| 34 wks to birth (cm/week)* | 0.64 (0.61,0.67) | 0.05 (0.004,0.09) | 0.69 (0.66,0.72) | -0.06 (-0.10,-0.01) |
| Birth (cm) | 36.62 (36.46,36.78) | 0.14 (-0.08,0.37) | 37.07 (36.91,37.22) | -0.75 (-0.97,-0.53) |
| Birth to 6 months (cm/week)* | 0.33 (0.32,0.35) | -0.01 (-0.03,0.003) | 0.34 (0.33,0.35) | -0.03 (-0.04,-0.01) |
| 6 months to 2 years (cm/week)* | 0.06 (0.05,0.06) | 0.004 (-0.001,0.01) | 0.06 (0.05,0.06) | 0.005 (-0.0003,0.009) |
| 2 years to 5 years (cm/week)* | 0.01 (0.01,0.01) | 0.001 (-0.001,0.003) | 0.01 (0.01,0.01) | 0.001 (-0.001,0.003) |
| 5 years (cm) | 51.91 (51.74,52.08) | 0.27 (0.03,0.51) | 52.50 (52.33,52.67) | -0.91 (-1.14,-0.68) |
| **Weight** |  |  |  |  |
| 20 weeks (kg) | 0.40 (0.39,0.42) | 0.002 (-0.02,0.02) | 0.41 (0.39,0.42) | 0.002 (-0.02,0.02) |
| 20 wks to 34 wks (kg/week)* | 0.16 (0.16,0.17) | -0.002 (-0.01,0.001) | 0.16 (0.16,0.17) | -0.002 (-0.01,0.002) |
| 34 wks to birth (kg/week)* | 0.24 (0.24,0.25) | 0.01 (-0.003,0.02) | 0.26 (0.25,0.26) | -0.02 (-0.03,-0.01) |
| Birth (kg) | 4.16 (4.11,4.21) | 0.01 (-0.06,0.08) | 4.24 (4.19,4.28) | -0.15 (-0.21,-0.08) |
| Birth to 6 months (kg/week)* | 0.17 (0.17,0.18) | -0.01 (-0.02,-0.001) | 0.18 (0.17,0.19) | -0.02 (-0.04,-0.01) |
| 6 months to 2 years (kg/week)* | 0.05 (0.05,0.06) | 0.005 (0.001,0.01) | 0.05 (0.05,0.06) | 0.004 (-0.0004,0.009) |
| 2 years to 5 years (kg/week)* | 0.04 (0.04,0.05) | 0.0004 (-0.002,0.003) | 0.04 (0.04,0.05) | -0.0003 (-0.002,0.002) |
| 5 years (kg) | 19.75 (19.43,20.08) | 0.15 (-0.31,0.61) | 20.08 (19.75,20.41) | -0.50 (-0.96,-0.05) |
| **Length/height** |  |  |  |  |
| Birth (cm) | 52.81 (52.56,53.06) | -0.13 (-0.48,0.22) | 53.16 (52.91,53.40) | -0.83 (-1.17,-0.48) |
| Birth to 6 months (cm/month)* | 0.66 (0.64,0.68) | -0.02 (-0.04,0.01) | 0.68 (0.66,0.70) | -0.06 (-0.08,-0.03) |
| 6 months to 2 years (cm/month)* | 0.24 (0.24,0.25) | 0.01 (0.001,0.02) | 0.24 (0.23,0.25) | 0.01 (0.01,0.02) |
| 2 years to 5 years (cm/month)* | 0.13 (0.13,0.14) | 0.0003 (-0.004,0.005) | 0.13 (0.13,0.14) | -0.0003 (-0.005,0.004) |
| 5 years (cm) | 109.72 (109.17,110.28) | 0.25 (-0.54,1.04) | 110.46 (109.90,111.03) | -1.22 (-2.01,-0.43) |

\*Change in growth per week/per month.

**Figure 1.**
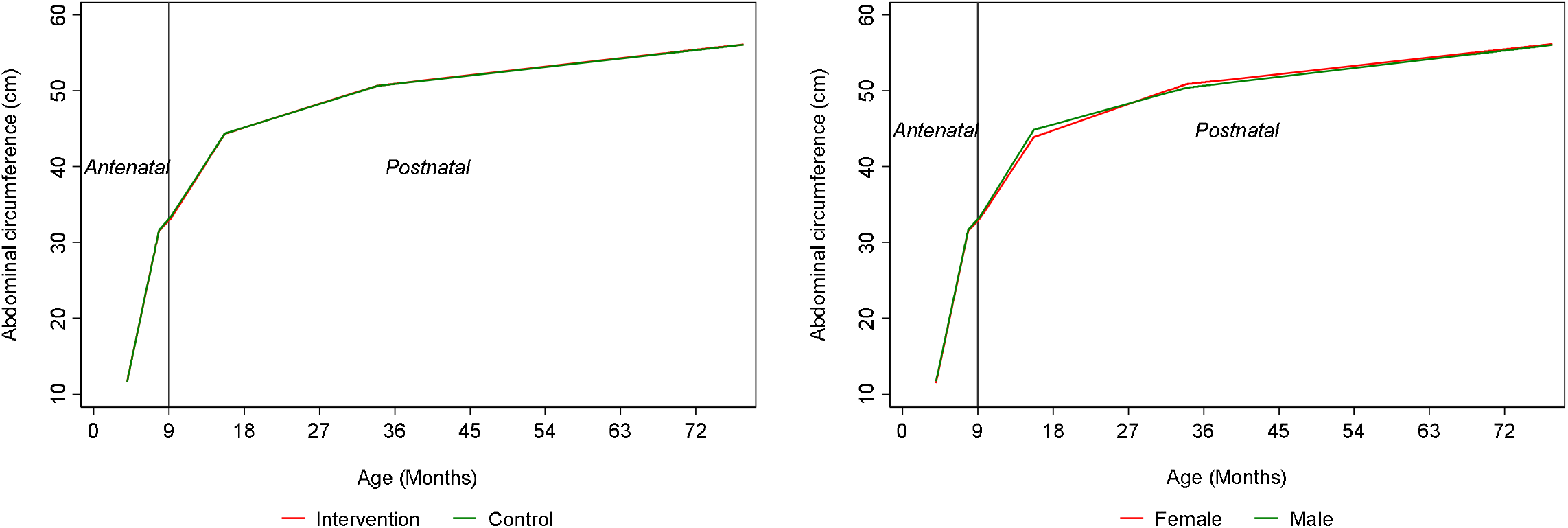
Trajectories of abdominal circumference from 20 weeks gestation to age five years by intervention status and sex. **Legend:** Note that X axis displays time in months because trajectory spans the antenatal and postnatal period.

**Figure 2.**
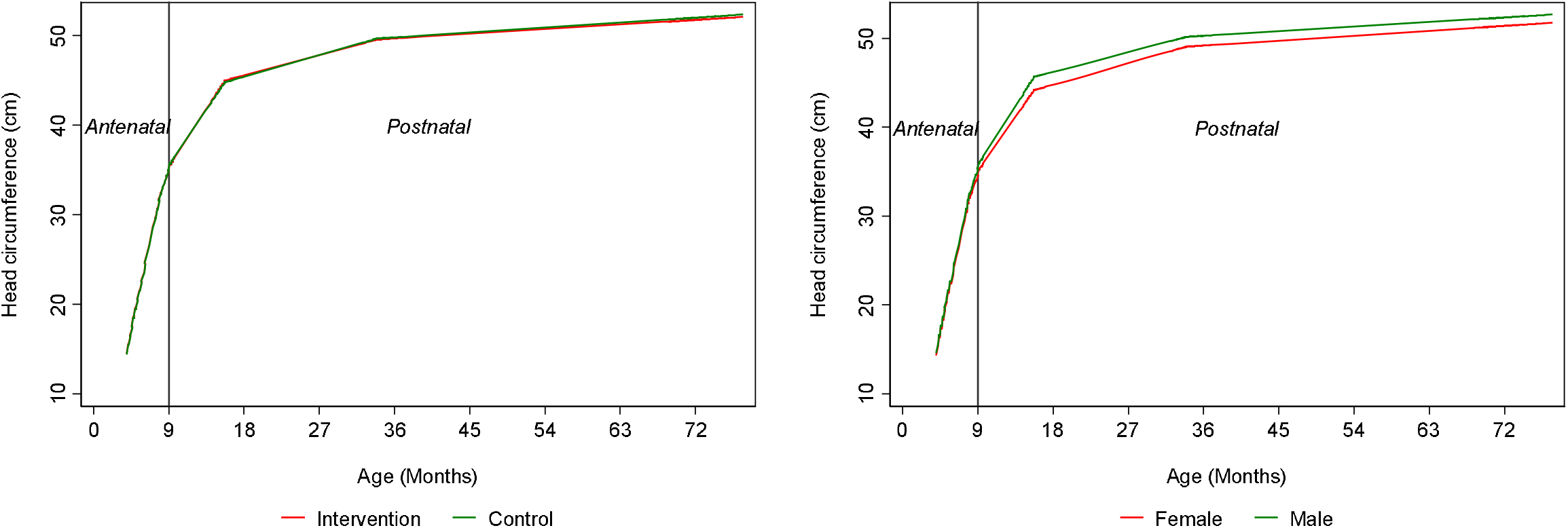
Trajectories of head circumference from 20 weeks gestation to age five years by intervention status and sex. **Legend:** Note that X axis displays time in months because trajectory spans the antenatal and postnatal period.

**Figure 3.**
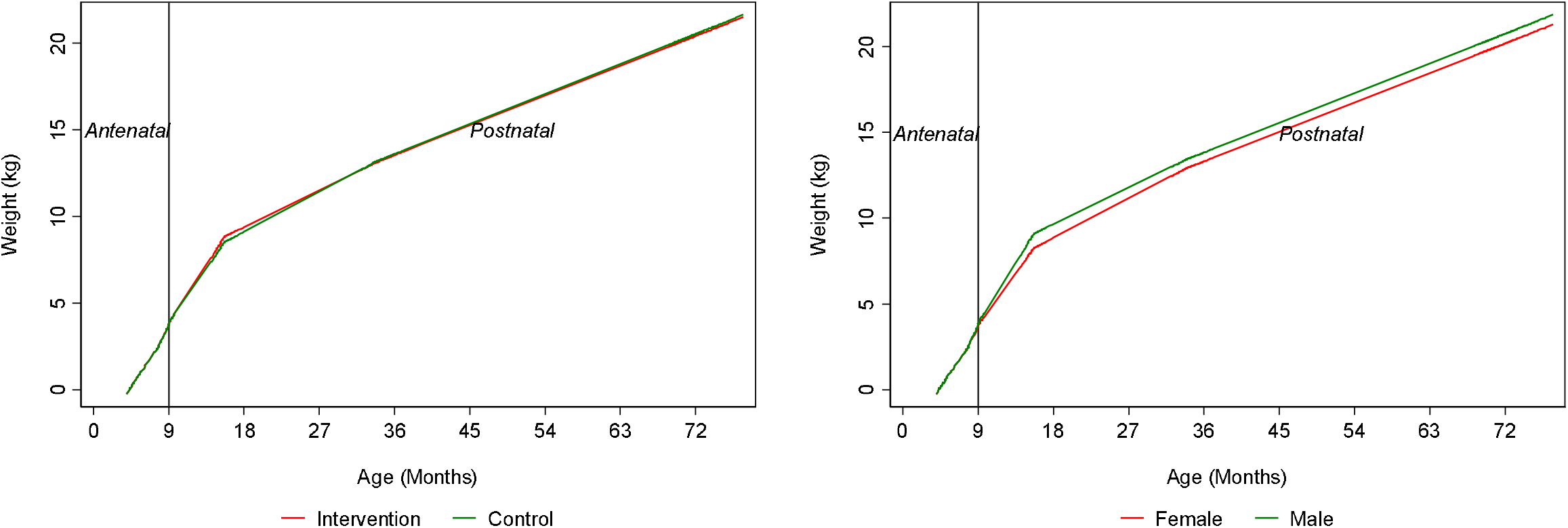
Trajectories of weight from 20 weeks gestation to age five years by intervention status and sex. **Legend:** Note that X axis displays time in months because trajectory spans the antenatal and postnatal period.

**Figure 4.**
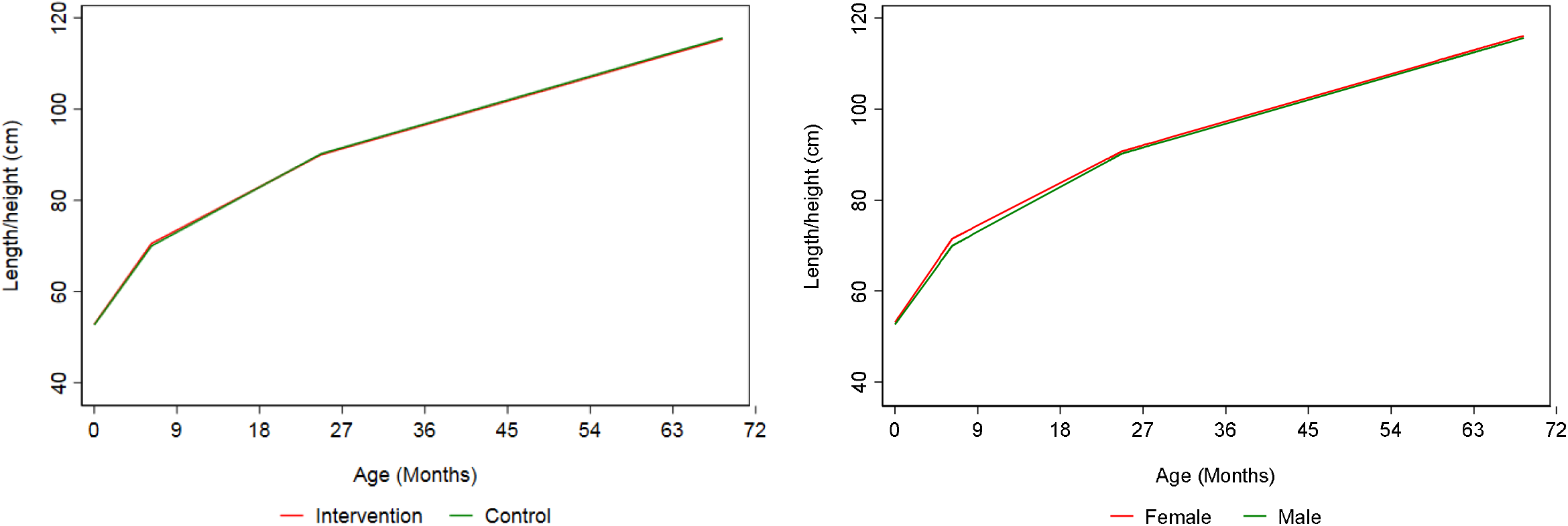
Trajectories of length/height from birth to age five years by intervention status and sex. **Legend:** Note that X axis displays time in months.

We found no strong evidence of differences in trajectories of AC, weight and length/height between the intervention and control group, but we found some evidence of slightly greater HC (difference 0.27 cm (95% Confidence Interval [CI] = 0.03, 0.51) emerging among the control group at five years. AC trajectories did not differ between males and females, though we found some evidence of modest differences in HC, weight and length/height trajectories between males and females. Females had lower HC at 20 weeks gestation with this difference widening at birth and persisting at age five years (difference at five years: - 0.91cm, 95% CI = -1.14, -0.68). Females had -0.15kg (95% CI = -0.21, -0.08) lower birth weight and slower postnatal growth rates in weight leading to -0.50 kg (95% CI = -0.96, - 0.05) lower weight among females at five years. Similarly, females were -0.83 cm (95% CI = - 1.17, -0.48) shorter in length at birth and had slower postnatal growth rates in length/height leading to -1.22 cm (95% CI = -2.01, -0.43) shorter height among females at five years.

## Discussion

In this prospective follow-up of a randomised control trial of approximately 750 infants at high risk of macrosomia, we demonstrated the use of linear spline multilevel modelling to examine trajectories of AC, HC, weight and length/height from 20 weeks’ gestation to age five years. We showed their applicability to data with repeated measures of growth which span the antenatal and postnatal period, even when as few as four repeat assessments are available (in the case of length/height) and measures are sparse. This work may be of value to other studies including randomised control trials with follow-up data such as ours in demonstrating the application of a multilevel modelling approach to examine growth trajectories which can subsequently be used as exposures or outcomes to better understand determinants and outcomes of growth in early life.

There are several strengths to the approach used here including ability to maximise sample sizes for analyses and reduce selection bias compared with traditional Z-score approaches since multilevel models can include all participants with at least one growth period under a MAR assumption (3). This is particularly advantageous where attrition rates from cohorts are high. Further advantages include more precise standard errors which consider the non-independence of repeated measures and here we have shown that the approach is implementable with as little as four repeated measures and with repeated measures that span antenatal and postnatal life. Limitations of this work include an inability to explore other non-linear growth patterns such as fractional polynomials due to the sparsity of measures which did not allow a range of possible shapes of growth trajectories to be explored (3). In cohorts with greater numbers of repeated measures and density of repeats, linear spline multilevel modelling can be implemented and compared to other possible shapes include fractional polynomials which have been shown to provide a more biologically intuitive shape of change(3). However, the linear spline approach demonstrated here provides many practical advantages including being more easily interpretable, allowing analysts to split trajectories into distinct periods of change that can then be easily related to exposures and outcomes. It should be noted that this cohort are unlikely to represent the growth rates or trajectories of a general population since their development is above average compared to what would be expected from an age and gender matched general population (the cohort is roughly approximated to the 75^th^ centile based on a crude comparison of means and SDs on the UK-WHO (Ireland) chart) (23).

## Conclusion

We demonstrate the application of multilevel linear spline models for examining growth trajectories when both antenatal and postnatal measures of growth are available. The approach may be useful for cohort studies or randomised control trials with repeat prospective assessments of fetal growth spanning pregnancy and childhood.

## Data Availability

All data produced in the present study are available upon reasonable request to the authors.

## Acknowledgements

The authors would like to thank all the ROLO participants for their involvement and all the staff of the National Maternity Hospital and the Perinatal Research Centre.

## Supplementary Material

### Sample code for implementing linear spline multilevel models using “runmlwin” command

This syntax utilises the user-written command ‘runmlwin’ which must be installed prior to use. The most recent version of MLwiN must be installed to be able to use this command and this package is available for use within Stata and R. Below we demonstrate the basic steps involved in implementing linear spline multilevel modelling suing “runmlwin” in Stata. Code below assumes data are in long format and that a variable called “occasion” exists identifying the ordering of observations within individuals. Sample code below applies to length/height from birth to five years.

### Generate the spline variable

First, three new variables are created: s1 (spline 1 from birth to 6 months), s2 (6 months to 2 year), s3 (2 years to 5 years).

mkspline s1_birth_6m 27 s2_6m_2 107 s3_2_max = age_lw

### Generate a constant term

MLwiN does not automatically include a constant term, so this must be generated and included in models.

gen cons=1

### Identify the location of MLwiN

global MLwiN_path “C:\Program Files\MLwiN v3.05\mlwin.exe”

### Run the multilevel model, sorting the data by person and occasion/age first

sort study_id age

runmlwin length cons s1_birth_6m s2_6m_2 s3_2_max ///

level2 (study_id: cons s1_birth_6m s2_6m_2 s3_2_max, reset(var) residuals (res, var)) ///

level1 (occ: age_lw, reset(var) diag) nopause maxiterations(150)

### Adding covariates

The following assumes covariates are binary and coded 0 and 1 or for covariates with multiple categories, dummy variables have been created. The addition of continuous covariates should be undertaken in the same manner as for categorical covariates but continuous covariates should be centred on the mean so that the baseline trajectory in the model is for the individuals with the mean level of the continuous covariate. Here we demonstrate the steps required for addition of sex as a covariate.

### Multiply covariate by splines

Once the covariate is coded in the format of 0/1 representing 0 for the baseline category, we multiply the covariate by the splines, creating interaction terms for inclusion in our model.

gen s1_birth_6m_fem = s1_birth_6m*female

gen s2_6m_2_fem = s2_6m_2*female

gen s3_2_max_fem = s3_2_max*female

### Run model now including covariate terms

The model is then ran as before but this time including a term for the covariate in question, here “female” and each of the above female*spline interaction terms generated. This allows the mean trajectory to differ for females and males. Because in this example the variable female is coded 0 for male and 1 for female the baseline trajectory is now for males with coefficients for “female”, s1_birth_6m_fem, s2_6m_2_fem, s3_2_max_fem representing the difference in the intercept, spline 1 and spline 2 and spline 3 in females compared with males.

sort study_id age

runmlwin length cons s1_birth_6m s2_6m_2 s3_2_max female2*, ///

level2 (study_id: cons s1_birth_6m s2_6m_2 s3_2_max, reset(var) residuals (res, var)) ///

level1 (occ: age_lw, reset(var) diag) nopause maxiterations(150)

## Notes

**Funding:** LMOK and KNON are supported by a Health Research Board of Ireland Emerging Investigator Award (EIA-FA-2019-007 SCaRLeT). The ROLO study was funded by the Health Research Board of Ireland, Health Research Centre for Health and Diet Research, and the European Union’s Seventh Framework Programme (FP7/2007-2013), project Early Nutrition under grant agreement no. 289346.

**Disclosures:** None of the authors have any conflicts of interest to declare.

### Competing Interest Statement

The authors have declared no competing interest.

### Funding Statement

LMOK and KNON are supported by a Health Research Board of Ireland Emerging Investigator Award (EIA-FA-2019-007 SCaRLeT). The ROLO study was funded by the Health Research Board of Ireland, Health Research Centre for Health and Diet Research, and the European Unions Seventh Framework Programme (FP7/2007-2013), project Early Nutrition under grant agreement no. 289346.

### Author Declarations

Institutional ethical approval and maternal written consent was received and carried out at the National Maternity Hospital, Dublin, Ireland.

